# Mutation Spectrum and Associated Risks of Medullary Thyroid Cancer and All-Cause Mortality in Incidentally Identified MEN2A-Causing *RET* Variants

**DOI:** 10.1101/2024.11.22.24317783

**Authors:** Courtney E. West, Uyenlinh L. Mirshahi, Katherine S. Ruth, Luke N. Sharp, Ankit M. Arni, Clare Turnbull, Caroline F. Wright, Bijay Vaidya, Martina M. Owens, David J. Carey, Kashyap A. Patel

**Affiliations:** Department of Clinical and Biomedical Sciences, University of Exeter Medical School, Exeter, UK; Department of Genomic Health, Geisinger, Danville, PA, USA; Institute of Cancer Research, London, UK; Department of Endocrinology, The Royal Devon University Healthcare NHS Foundation Trust, Exeter, UK; The Exeter Genomics Laboratory, The Royal Devon University Healthcare NHS Foundation Trust, Exeter, UK

## Abstract

**Importance:** *RET* pathogenic variants cause Multiple Endocrine Neoplasia type 2 (MEN2), characterised by medullary thyroid cancer (MTC). With increasing incidental identification of these variants in asymptomatic individuals outside family screening, their risk of MTC and all-cause mortality without intervention remain unknown in this context.

**Objective:** To determine the risk of MTC and all-cause mortality in clinically unselected individuals and assess how the risk of MTC differ from clinically ascertained cases.

**Design, Setting, and Participants:** Prospective cohort study of 383,914 unrelated individuals from the clinically unselected UK population (UK Biobank) and 122,640 from the US health system (Geisinger cohort). We compared MTC risk in these cohorts to 1,078 individuals who were clinically ascertained with suspicion of MEN2 from UK routine practice.

**Exposures:** *RET* pathogenic variants causing MEN2

**Main Outcomes and Measures:** Frequency and the spectrum of pathogenic *RET* variants, Risk of clinically presented MTC, all-cause mortality without thyroidectomy.

**Results:** Pathogenic *RET* variants were found in 0.04% of individuals from UK population cohort and 0.08% of individuals from US health system cohort. They were predominantly from moderate-risk category as per American Thyroid Association guideline (99.4% and 94.8% respectively). MTC risk by age 75 in variant carriers in the UK population was 2.2% (95% CI 0.7-6.8) and 19% (95% CI 5.7-30) in US health system cohort. This was significantly lower than the clinically ascertained cohort with the matched variants (95.7%, 95% CI 82.1-99.7 p<0.0001). In the UK Biobank, most variant carriers (98.2%) did not undergo thyroidectomy and their all-cause mortality by age 75 was similar to non-carriers (6.1%, 95% CI 2.7-13.8 vs 5.7%, 5.6-5.8, p=0.79), with consistent findings in the US health system cohort.

**Conclusions and Relevance:** Moderate-risk *RET* variants are most common in incidental cases. These variants carry substantially lower MTC risk than clinically ascertained cases. This evidence addresses a current knowledge gap, enabling more informed clinical decision-making.

## Introduction

Gain-of-function pathogenic variants in the *RET* oncogene cause Multiple Endocrine Neoplasia type 2 (MEN2), an aggressive autosomal dominant endocrine cancer syndrome.^1,2^ Medullary thyroid cancer (MTC) is the most common feature, affecting 95-100% of patients with MEN2^1,2^ Genetic testing of *RET* is widely recommended for all cases with MTC and/or suspected MEN2.3 If a patient is found to have a pathogenic *RET* variant, cascade testing of their relatives is performed and heterozygous relatives considered for early curative prophylactic total thyroidectomy due to high risk of MTC^2–4^

Current guidelines recommend reporting incidentally identified pathogenic *RET* variants in asymptomatic individuals.5, 6 These are the individuals where a pathogenic *RET* variant is identified as part of secondary findings outside of the family screening. This could be identified via diagnostic clinical exome/genome sequencing for unrelated indications or as part of research studies (clinically unselected cases). Since 2013, when the American College of Medical Genetics and Genomics (ACMG) first recommended this approach for MEN2A-causing *RET* variants, ^5^ it has been widely emulated including by the UK’s 100,000 Genomes Project, European Society for Medical Oncology,^6^ and recent newborn genomic screening proposals.^7,8^ The presumption of net clinical benefit that underpins these recommendations has been extrapolated from the benefit-risk balance ascribed to clinically identified families, on account of the lack of data in incidentally detected cases. This difference in approach warrants careful evaluation, particularly given the distinct contexts in which these variants are being identified. Recent studies have demonstrated that analyses of clinically ascertained families tend to overestimate risk for monogenic disorders compared to those whose pathogenic variants were identified through incidental findings. ^9,10^ Consequently, determining the frequency, mutational spectrum, rate of clinical MTC presentation in incidental *RET* variant carriers, and understanding how this differs from clinically ascertained cohorts, is important to inform the current recommendations for incidentally identified cases. This knowledge gap has become increasingly pressing with the widespread adoption of exome/genome sequencing in clinical and research settings, coupled with recommendations to report incidental *RET* findings, highlighting the urgent need to better understand MTC risk in incidental cases.

We therefore aimed to identify frequency of pathogenic *RET* variants, the spectrum of variants and their associated risks of MTC and all-cause mortality without intervention in clinically unselected individuals. We analysed detailed genetic and clinical data from over 500,000 clinically unselected individuals who underwent genomic sequencing. We also compared the risk of clinically presented MTC in these individuals to clinically ascertained cases to highlight the differences between these two groups.

## Methods

### Study Populations

#### UK Biobank

UK Biobank is a large UK population cohort of 500,000 clinically unselected individuals recruited between 2006-2010 at ages 40-69 years. ^11^ It contains phenotypic data provided through self-reported questionnaires, hospital records, cancer, and death registries, and linked general practitioner (GP) records available both at baseline and follow-up.^11^ We utilised 383,914 individuals from exome sequencing data release on 450,000 individuals who were unrelated (up to third degree). ^12^ We used unrelated individuals to prevent undue influence of large families on variant frequency in the cohort (15.6% related). We used linked data on phenotypes at baseline and follow up (up to October 2022). Baseline characteristics of this cohort are summarised in **Supplementary Table 1**. The UK Biobank Research Ethics Committee approved the study, and all participants provided written informed consent.

**Table 1:** Pathogenic medullary thyroid cancer-causing *RET* variants in the unrelated individuals in clinically unselected study cohorts.

|  |  | Population cohort<br>UK Biobank<br>(unrelated) | Health care system<br>cohort, USA<br>Geisinger MyCode<br>(unrelated) |
| --- | --- | --- | --- |
| **All *RET* pathogenic variant carriers** |  | 169/383,914 (0.04%) | 77/122,640 (0.06%) |
| **Age at recruitment, y** |  | 57.0 (8.09) | 56.2 (17.8) |
| **Female sex** |  | 75 (44.4) | 45 (58.4) |
| **Pathogenic Variant classification** |  |  |  |
| *Highest risk* | p.(Met918Thr) | 0 | 0 |
| *High risk* | P.(Cys634Arg/Phe/Trp/Tyr)<br>p.(Ala833Phe) | 1 (0.5%) | 4 (5.2%) |
| *Moderate risk* | All | 168 (99.5%) | 73 (94.8%) |
|  | p.(Cys609Phe/Tyr)*or<br>p.(Cys620Gly/Trp/Tyr)*<br>or.(Asp631Tyr)* | 8 | 8 |
|  | p.(Lys666Asn) | 14 | 6 |
|  | p.(Lys666Glu) | 6 | 2 |
|  | p.(Glu768Asp) or p.(Ser904Phe)<br>or p.(Met918Val) | 7 | 2 |
|  | p.(Leu790Phe) | 14 | 0 |
|  | p.(Val804Met) | 95 | 30 |
|  | p.(Val804Leu) | 15 | 0 |
|  | p.(Ser891Ala) | 9 | 25 |

Pathogenic variants grouped according to American Thyroid Association Guideline (3). Number of carriers with each variant is indicated. Mean (SD) for continuous data and n (%) for categorical data. \*Extracellular moderate risk variants. Variants with n<5 were combined in line with UK Biobank policy

#### Geisinger MyCode Cohort

The Geisinger MyCode cohort is a health system-based cohort from the USA consisting of 340,423 unselected individuals who sought care at Geisinger, a healthcare provider in central and northeastern Pennsylvania. Participants have consented to share their routinely collected electronic health records for research, including clinical diagnoses, procedures, cancer registry data, medications, and laboratory results. The cancer registry included histological data of cancers diagnosed and treated at Geisinger since 1943. We used a subset of 122,640 unrelated MyCode participants (up to third degree) with exome sequencing, generated as part of the DiscovEHR collaboration between Geisinger and the Regeneron Genetics Center.^14^ We used unrelated individuals to prevent undue influence of large families on variant frequency in the cohort (36% related up to third degree). We used data at baseline (the date of exome sequencing) and follow up (up to June 2023). Our study was reviewed by the Geisinger Institutional Review Board and determined not to be human subjects research as defined in 45CFR46.102(f) (Study #2016-0269). The cohort characteristics at completion of exome sequencing (baseline) are summarised in **Supplementary Table 1** and have been extensively described previously. ^9^

#### Exeter clinical cohort

This includes 1,078 unrelated individuals (probands) from routine clinical practice in the UK with suspected MEN2 who were referred to the Exeter Genomics Laboratory for *RET* gene sequencing from 1995-2018. Consent was provided by patients or parents prior to genetic testing, and part of the data from this cohort was previously published.^15^ Of the 1,078 individuals, 117 were found to have a heterozygous germline pathogenic activating *RET* variant. The baseline characteristics of the clinical cohort are summarised in **Supplementary Table 1**.

### Genetic Analysis

#### UK Biobank

We used exome sequencing data for our study. This was released centrally by the UK Biobank. The detailed process of exome sequencing and sample/variant filtering have been described by Szustakowski et al. ^11^ and are available at https://biobank.ctsu.ox.ac.uk/showcase/label.cgi?id=170].

#### Geisinger MyCode Cohort

We used exome sequencing in our study. This was performed as part of the DiscovEHR collaboration between Geisinger (Danville, PA) and the Regeneron Genetics Center (Tarrytown, NY). ^15^ The detailed method for exome sequencing and sample/variant filtering has been described previously by Mirshahi et al. ^9^

#### Exeter clinical cohort

We used sanger sequencing or targeted gene panels for clinical *RET* sequencing. Sequencing coverage included but not limited to exons 10, 11, 13, 14, and 16 of *RET*. ^15^ Clinical scientists at the Exeter Genomics Laboratory, the Royal Devon and Exeter Hospital analysed the variants as part of routine diagnostic care.

This laboratory has provided *RET* genetic testing for the UK population since 1995 and considered a centre of excellence for *RET* genetic testing in the UK.

### Variant classifications in clinically unselected cohorts

We annotated *RET* variants using MANE Select transcript NM_020975.5. We reviewed all heterozygous missense variants in *RET* across the two clinically unselected cohorts. Variants were classified as pathogenic if previously reported in probands with MEN2 and deemed likely pathogenic or pathogenic according to the American College of Medical Genetics and Genomics/the Association for Molecular Pathology (ACMG/AMP) guidelines^5^ in conjunction with the ACGS Best Practice Guidelines for Variant Classification in Rare Disease. ^16^ We utilised three *RET* pathogenic variant databases and the ClinVar database to check for previous reports of the variants in patients with MEN2. ^15,17–19^ We manually reviewed the sequence reads supporting pathogenic variants using the Integrative Genomics Viewer (IGV), assessing read depth and allele fraction to remove false positives or somatic mosaics. All pathogenic variants were considered high quality based on two independent reviews. We also classified variants into highest, high, or moderate risk categories based on American Thyroid Association (ATA) recommendation.^3^

### Phenotype definitions

#### Clinically unselected cohorts

We used two definitions to identify clinically presented MTC cases. Our main definition included malignant thyroid cancer with medullary histology, whilst our broad definition additionally included any thyroid cancer or thyroidectomy (partial/subtotal/total) regardless of indication. This broader definition captured cases without cancer registry data, missing histology, or those undergoing prophylactic surgery. We identified cases using hospital episodes statistics (ICD10), operation records (OPCS), cancer registry, death registry, GP records and self-reported data. The main definition served as our primary analysis, with the broad definition used for sensitivity analysis. Data sources and codes for both definitions are detailed in **Supplementary Table 2**.

All data were collected up to the date of recruitment (UK biobank) and follow up to October 2022. For Geisinger cohort, we used data up to the date of exome sequencing. The follow up data excluded individuals whose incidental findings pathogenic *RET* variants were reported back to them as part of MyCode Genome Screening and Counselling Program that discloses results to biobank participants. The findings for these individuals were recently published elsewhere. ^20^ For individuals where incidental findings have not yet been reported, we presented follow up data from exome sequencing to 12 June 2023. We used death registration to assess all-cause mortality from recruitment in all clinically unselected cohorts.

#### Exeter Clinical cohort

The referring clinician reported the presence/absence of MTC and age at MTC diagnosis at the time of referral to Exeter Genomics Laboratory for genetic testing. Where this was not explicitly stated, the age of diagnosis was taken as the age of referral for analysis of the *RET* gene.

### Statistical analysis

We assessed penetrance by calculating the proportion of MTC cases among carriers with *RET* pathogenic variants. We used exact binomial 95% confidence intervals for the proportions. We calculated age-related penetrance using Kaplan-Meier survival analysis, given the availability of age at diagnosis/surgery information. We followed participants until study end (October 2022 for UK Biobank and June 2023 for Geisinger cohort), censoring at disease onset (using main or broad definition), or death. We analysed MTC incidence rates and all-cause mortality from recruitment to study end in both cohorts. We used log-rank tests for equality to compare age-dependent penetrance between groups and computed Cox proportional hazard ratio adjusting for age at recruitment and sex. We used Stata 16 (College Station, Texas, USA) and the survminer 0.4.9 package in R (R Foundation for Statistical Computing) for data analysis.

## Results

### The prevalence of MEN2-causing pathogenic *RET* variants was 0.04-0.06% in clinically unselected cohorts, of which the majority were moderate risk variants

Analysis of 383,914 unrelated individuals with exome data in the UK Biobank, a population cohort from the UK, identified 169 carriers of one of 17 different MEN2-causing pathogenic *RET* variants, giving a prevalence of 1 in 2500 (0.04%, 95% CI 0.038-0.051%) (Table 1). Of these 17 variants, 16 (94%) were moderate risk (based on the American Thyroid Association guideline) and present in 168 individuals, and one individual carried a high-risk variant (Table 1, Supplementary Table 3). The most common variant was p.(Val804Met), found in 95 individuals. (Table 1) The prevalence of pathogenic *RET* variants in the large US health system-based Geisinger cohort was slightly higher at 1 in 1666 (0.06%, 95% CI 0.05-0.78%), likely attributable to the hospital-based setting which would include relatively more individual with self-presented MEN2. Aligning with the UK Biobank findings, 71% (10/14) of variants were moderate risk with p.(Val804Met) being most common and observed in 39% (30/77) *RET-*harbouring individuals, while 28% (4 variants) were high risk observed in 5% (4/77) of individuals (Table 1).

### Low MTC risk in *RET* pathogenic variant carriers from UK population cohort

Among 168 *RET* pathogenic variant carriers in UK Biobank, only two (1.2%, 95% CI 0.1-4.2%) had MTC at recruitment, one individual with a high risk variant and one with a moderate risk variant (Supplementary Table 4). After 2,299 person-years of follow-up (median follow up 13.8 years (IQR 13.0-14.4) at average age 70.6 years (SD 8.1), we identified one additional case (0.41 cases per 1,000 person-years, 95% CI 0.11-2.35). Overall MTC penetrance was 2.2% (95% CI 0.7-6.8%) by age 75 years (Figure 1A, 1B). Although the absolute risk was low, it was still relatively higher in *RET* carriers compared to non-carriers (age and sex adjusted hazard ratios 334, 95% CI 99.2-1125, p<0.0001). Using a broader definition that included any thyroid cancer or thyroidectomy identified two additional cases, increasing penetrance to 2.78% (95% CI 1.0-7.4%) by age 75 (Figure 1A, 1C; Supplementary Table 4). In line with the broader less specific definition, HR was lower, 2.84 (95% CI 1.1-7.6, p=0.037). All three additional cases had thyroidectomy for non-MTC indications (Supplementary Table 4). The penetrance was similar between p.(Val804Met) and other moderate-risk variants (p=0.84) and also intracellular and extracellular moderate risk variants (p=0.75). The results by variant risk categories are provided in Supplementary Table 5.

**Figure 1:**
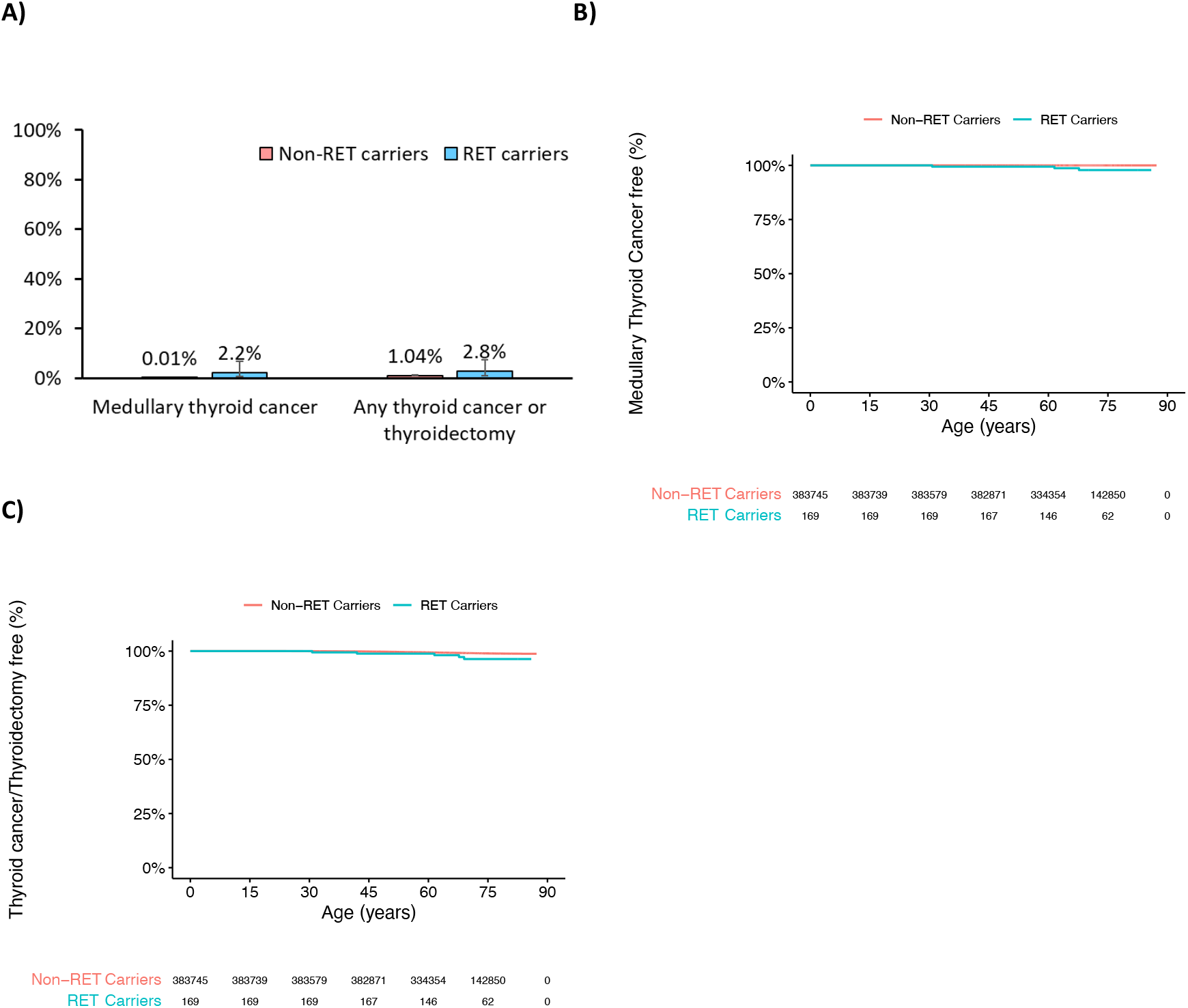
The age-related risk of medullary thyroid cancer in individuals with pathogenic *RET* variants is very low in a clinically unselected UK Biobank cohort. **A)** Bar Graph showing propor.on of unrelated individuals in the UK Biobank with prevalent and incident medullary thyroid cancer by age 75 years, and with any thyroid cancer or any thyroidectomy (par.al/total/subtotal) for any indica.on, by *RET* pathogenic variant status (n=169 carriers and 383,745 non-carriers). The error bar represents 95% confidence interval. **B)** Kaplan-Meier plots showing age-related penetrance of MTC for both prevalent and incident cases, **C)** and for any thyroid cancer or thyroidectomy (broad defini.on), Log-rank test showed significant differences between RET carriers and non-carriers (p=3×10^−193^ for MTC; p=0.02 for broad defini.on) with age and sex adjusted hazard ra.os of 334 (95% CI 99-1125, p<0.0001) and 2.84 (95% CI 1.1-7.6, p=0.037), respec.vely

### Lower MTC risk in incidentally identified cases supported by replication in the independent US health system cohort

To validate our findings, we analysed independent 122,640 unrelated individuals from the US-based Geisinger MyCode cohort. Among 77 *RET* pathogenic variant carriers, MTC penetrance by age 75 was 19% (95% CI 5.7-30%) (Figure 2A, 2B) and 24.3% (95% CI 10.7-35.9%) using a broad definition (Figure 2A, 2C). MTC penetrance was 25% (1/4, 95% CI 0.6-80.5%) in high-risk variant carriers and 12.3% (9/73, 95% CI 5.8-22.1%) in moderate-risk carriers (Supplementary Table 5). The overall age and sex adjusted hazard ratios for MTC in all variant carriers was 1261 (95% CI 545-2916, p<2×10^−16^) and for broad definition was 12.3 (95% CI 7.2-20.5, p<2×10^−16^). Of 77 cases, 28 yet uninformed of their *RET* mutation status as part of MyCode Genome Screening and Counselling Program that discloses results to biobank participants, and none of them developed MTC over 538 person-years of follow-up (median 7.0 years, IQR 4.4-8.9) at a median age of 74.5 years (IQR 58-84). Outcomes for remaining 49 participants informed through the MyCode Genome Screening and Counselling Programme were recently published^20^. MTC cases stratified by ATA risk categories and the common p.(Val804Met) variant are detailed in Supplementary Table 5.

**Figure 2:**
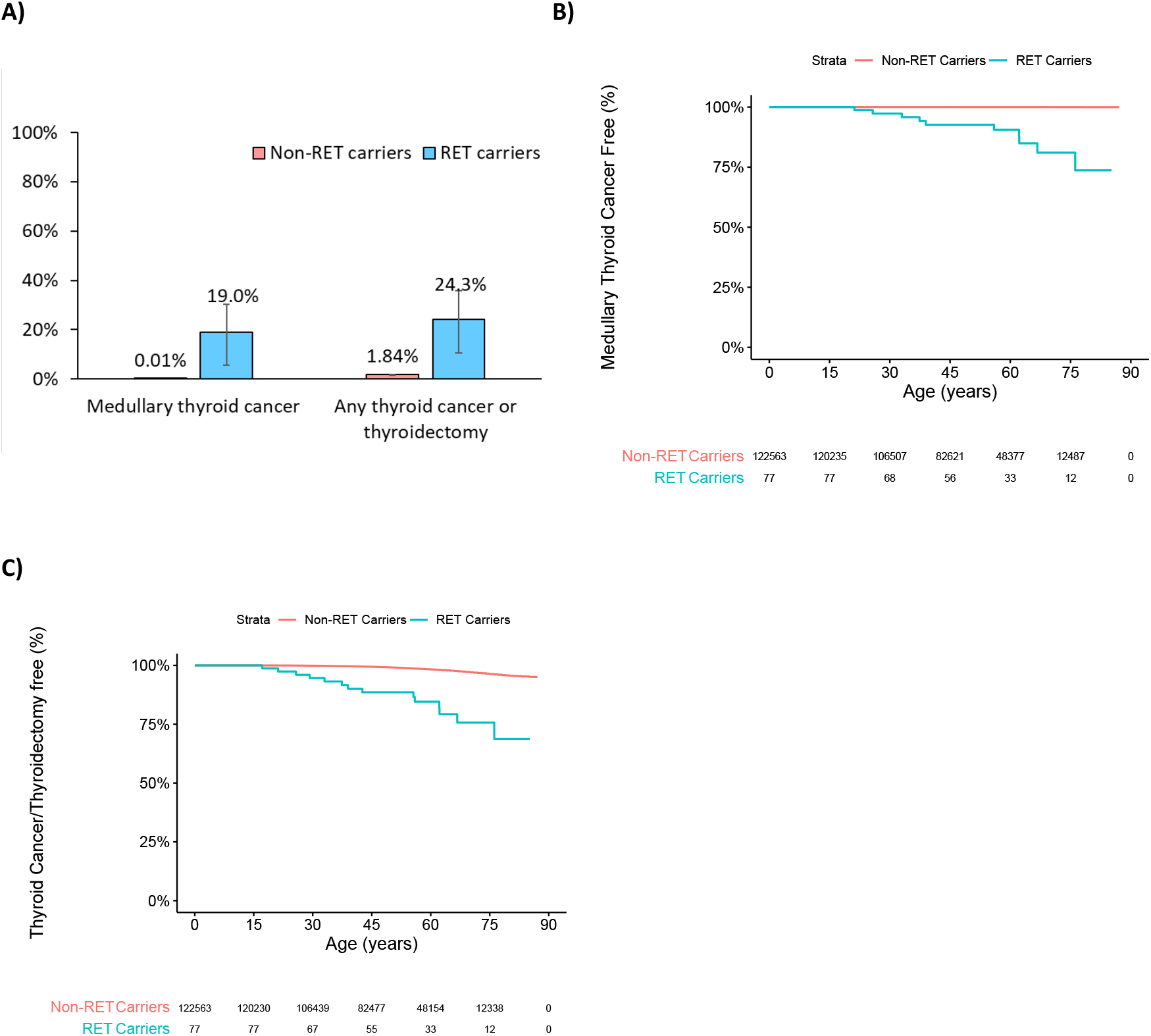
The lower risk of medullary thyroid cancer in clinically unselected individuals with pathogenic RET variants replicated in health system-based cohort. **A)** Bar Graph showing propor.on of unrelated individuals in the Geisinger health system cohort, Pennsylvania, USA with prevalent medullary thyroid cancer by age 75 years, and with any thyroid cancer or any thyroidectomy (par.al/total/subtotal) for any indica.on, in RET pathogenic variant carriers (n=77) and non-carriers (n=122,563). The error bar represents 95% confidence interval. **B)** Kaplan-Meier plots showing age-related penetrance of MTC for the prevalent cases, **C)** and for any thyroid cancer or thyroidectomy (broad defini.on), Log-rank test showed significant differences between RET carriers and non-carriers (p<2×10^−16^ for MTC; p<2×10^−16^ for broad defini.on) with age and sex adjusted hazard ra.os of 1261 (95% CI 545-2916, p<2×10^−16^) and 12.3 (95% CI 7.2-20.5, p<2×10^−16^), respec.vely.

### Incidentally identified pathogenic RET variant carriers did not show excess all-cause mortality without prophylactic thyroid surgery

A genotype-first approach uniquely enabled us to assess mortality outcomes in untreated *RET* variant carriers, previously impossible as withholding intervention was considered unethical, particularly in high-risk family members. In UK Biobank, 166 (98%) variant carriers had not undergone thyroidectomy. Over 2,299 person-years of follow-up (median 13.8 years, IQR 13.0-14.4), their all-cause mortality was comparable to non-carriers (6.1%, 95% CI 2.7-13.8 vs 5.7%, 5.6-5.8, by age 75 years, log-rank test p=0.79, HR = 0.94, 95% CI 0.42-2.1, p=0.88) (Figure 3A). Mortality rates were similar between carriers with and without thyroidectomy (p=1), though analysis was limited by few thyroidectomy cases. These findings were consistent in the Geisinger cohort, where none of the 28 uninformed *RET* variant carriers had undergone thyroidectomy over 140 person-years of follow-up (median 5.03 years, IQR 2.7-7.9) and showed no excess mortality compared to non-carriers by age 75 years (11.6%, 95% CI 0-21.8 vs 13.9%, 95% CI 13.5-14.2, log-rank test p=0.5, HR = 1.43, 95% CI 0.79-2.6, p=0.24)(Figure 3B).

**Figure 3:**
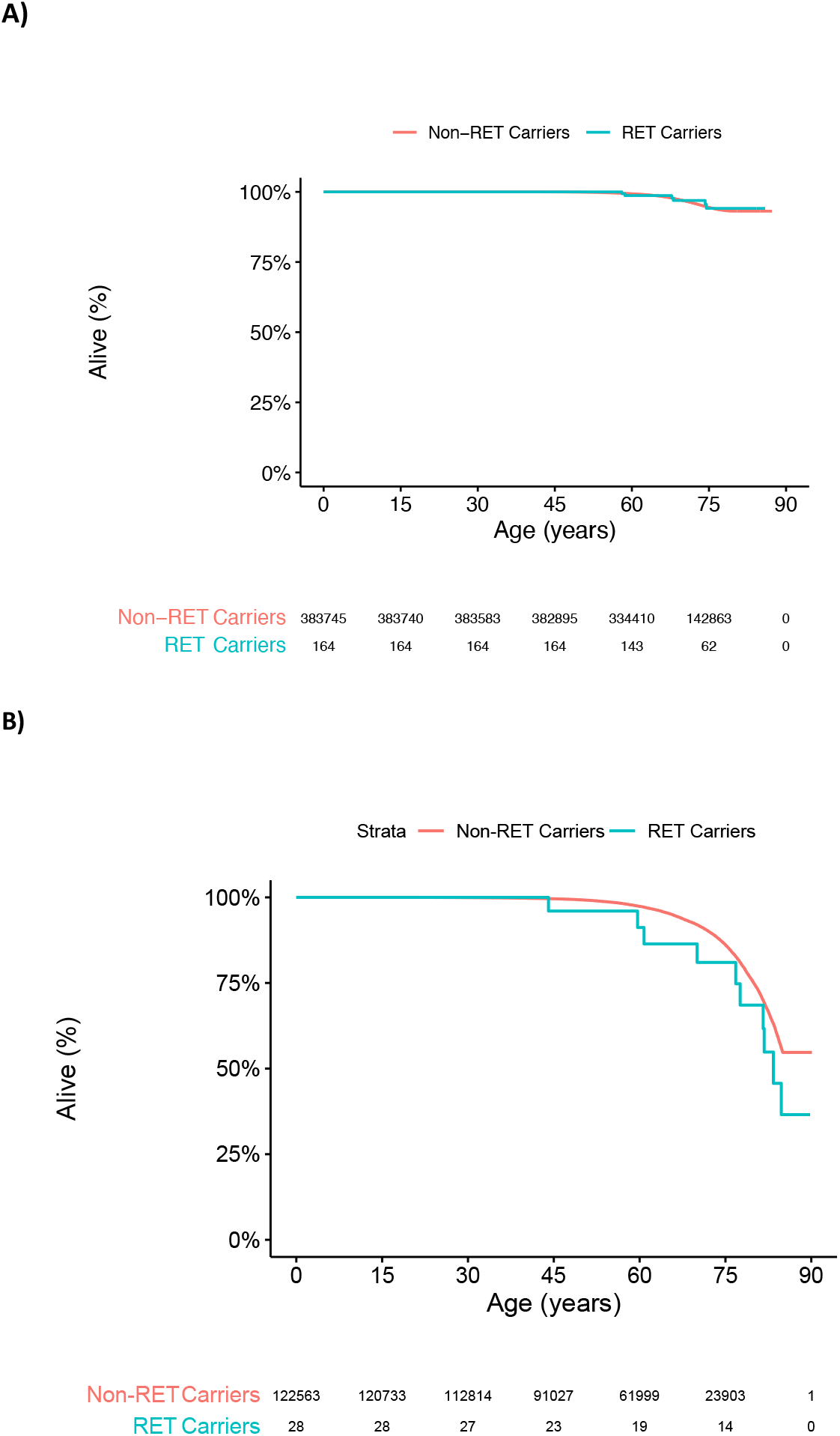
Pathogenic *RET* variant carriers did not show excess all-cause mortality without prophylactic thyroid surgery. All-cause mortality in *RET* pathogenic variant carriers and non-carriers without thyroidectomy for any indication in **A)** UK Biobank over median 13.8years follow up (log-rank test p=0.81, *age and sex adjusted* Hazard ratio = 0.94, 95% CI 0.42-2.1, p=0.88) **B)** Geisinger MyCode cohort over median 7.0 years follow up at a median age of 74.5 (log-rank test p=0.24, *age and sex adjusted* hazard ratios of 1.43, 95% CI 0.79-2.6).

### Incidentally identified *RET* variants show significantly lower MTC penetrance than clinical cases

We compared MTC penetrance between clinically unselected cases and 1,078 probands referred from UK routine clinical practice due to suspected MEN2. Of these clinical referrals, 117 (10.85%) carried a pathogenic *RET* variant (55% moderate-risk, 30% high-risk, 15% highest-risk). Age-dependent MTC penetrance in clinical referrals was 68% and 84% at ages 30 and 45 for highest-risk variants, 35% and 70% for high-risk variants, and 16% and 45% for moderate-risk variants, respectively (Figure 4A). When analysing only variants present across all three cohorts, MTC penetrance at age 75 was significantly higher in clinically ascertained cases (95.7%, 95% CI 82.1-99.7) compared to Geisinger (15.9%, 95% CI 8.0-30.2) and UK Biobank (1.32%, 95% CI 0.2-8.7), both p<10^−16^ (Figure 4B). This difference persisted when analysing only the most common variant p.(Val804Met) (92.3% vs 1.6% vs 0%) and other moderate-risk variants (Supplementary Figure 1). Similarly, for high-risk variants we observed directionally consistent results but limited by small numbers (40%, 2/5 vs 83%, 29/35)(Fisher’s exact test p=0.03; Supplementary Figure 1C, Supplementary Table 5).

**Figure 4:**
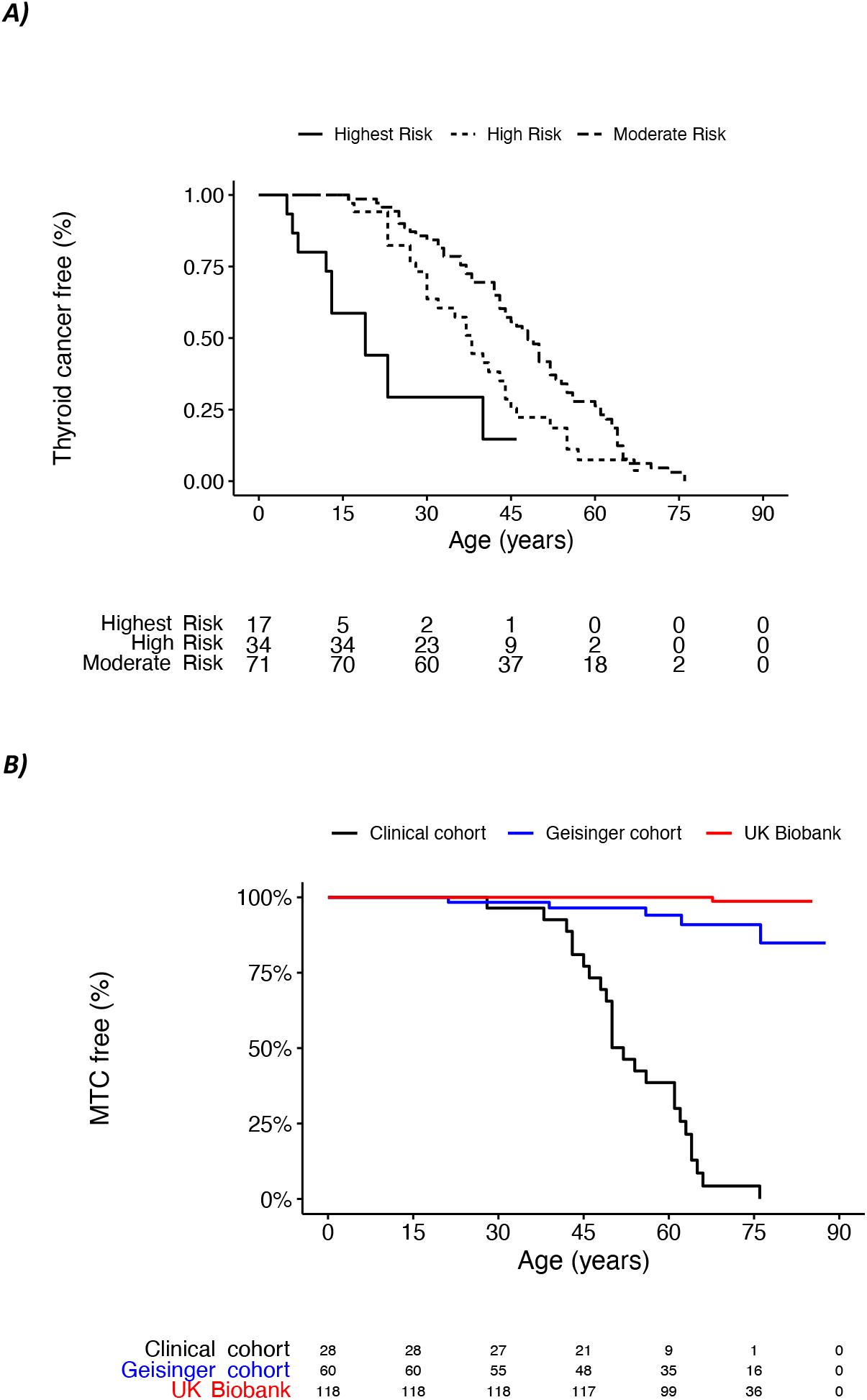
The age-related risk of medullary thyroid cancer in individuals with *RET* pathogenic variant ascertained clinically and in clinically unselected populaHon and health system-based cohort with the matched variants. A) Kaplan-Meier plot demonstra.ng the age-related penetrance of MTC for *RET* pathogenic variant carriers iden.fied from individuals referred to the Exeter genomic laboratory for gene.c tes.ng in rou.ne clinical prac.ce in the UK with suspected MEN2A. B) Kaplan-Meier plot illustra.ng the age-related penetrance of MTC for the matched variants across clinically selected and unselected popula.on cohort (UK Biobank) and health care system-based cohort (Geisinger cohort). Log rank test p<10^−16^ (clinical vs Geisinger) and p<10^−47^ (clinical vs UK Biobank).

## Discussion

Our genomic screening of nearly 500,000 unselected individuals from two different ascertainment settings suggests that incidental pathogenic activating *RET* variants occur in approximately 1 in 2,000 individuals— 30-fold more common than previously estimated^21^. The vast majority are moderate-risk variants, which associate with low MTC risk and does not show excess all-cause mortality without thyroidectomy.

Accurate MTC risk information is essential for effective risk-benefit discussions with individuals carrying incidentally identified pathogenic *RET* variants. While intervention choice remains personal, our study provides the first evidence in this context to support informed decision-making by healthcare professionals and genetic counsellor. We present penetrance values for different genotype-first settings, enabling counsellors to provide more appropriate risk estimates. We demonstrate that current risk estimates based on family studies overestimate the risk, a pattern now observed across several monogenic disorders including breast cancer, Lynch syndrome, and diabetes.^9,10,22^. Our findings primarily relate to moderate-risk variant carriers, as our sample size for high-risk variants was limited (n=5, two with MTC), highlighting the importance of variant-specific penetrance estimates as deleterious variants within the same gene can impart different risks for the same condition. The consistency of results with broad disease definition with longitudinal data, across two healthcare settings in two countries supports the validity of our results. Our p.(V804M) penetrance estimate aligns with Loveday et al’s prediction of 4% (95% CI 0.9-8%), based on maximum tolerated allele Frequency (MTAF)^23^ as well as lower estimate (3-20%) observed from family-based studies.^24^ The risk estimates for moderate risk variants excluding p.(Val804Met) were similar to other moderate risk variants (Supplementary table 5). Although extracellular moderate risk variants compared to intracellular variants showed higher MTC risk in Geisinger but not in UK Biobank carriers (Supplementary table 5). The higher penetrance at Geisinger likely reflects its healthcare-based recruitment which would include relatively more cases with self-presented MEN2, shown by its higher MTC prevalence (18.7/100,000) compared to previous population estimates (3.8/100,000). While UK Biobank’s MTC prevalence (7.5/100,000) aligns with population estimates, its healthy volunteer effect suggests our penetrance estimates likely represent a lower bound of true population penetrance 25,26

Our results contrast with a recent study by Pichardo *et al*, ^20^ which reported outcomes from 20 Geisinger MyCode participants undergoing prophylactic thyroidectomy after incidental detection of moderate-risk *RET* p.(Ser891Ala)(n=14) and p.(Val804Met)(n=6). Of 75 moderate-risk *RET* carriers offered surgery, only 20 (26%) proceeded, with 12 showing histologically confirmed MTC, predominantly stage one disease (10/12, 83.3%). Outcomes for those declining surgery were not reported. The absence of excess mortality and lower rate of clinically presented MTC in our cohort of incidentally identified carriers unaware of their *RET* status perhaps suggests that some early-stage disease found through screening thyroidectomy might not progress to clinically significant disease. The high prevalence of C-cell hyperplasia in both moderate-risk variant carriers and healthy individuals,^27^ coupled with the low MTC risk we observed, may support this hypothesis. Further research is needed to understand this lower MTC risk in incidentally identified cases and explore potential genetic modifiers of penetrance as previously reported in *BRCA*-related cancer and monogenic diabetes. ^9,10,28^

Our study addresses the previously missing evidence on incidentally identified pathogenic *RET* variants. Current recommendations are based on the benefits and risk observed in clinically ascertained families due to lack of available data for large numbers of incidentally identified cases. We present the first comprehensive data in this context, including disease prevalence (∼1:2000), variant distribution (∼98% moderate risk), and disease risk (2.2-16% at age 75) and all-cause mortality in the absence of prophylactic thyroidectomy (no impact). The surgical risks with prophylactic thyroidectomy although low (<10% chance of long-term complications)^29^ coupled with lifelong hormone dependence affecting quality of life in up to 20% of cases may become particularly pertinent against the lower MTC risk in incidentally identified moderate-risk variant carriers.^4,29,30^ Although variant reporting may continue and intervention choice remains personal, if one opts for biomarker-driven monitoring in this context, our data suggest this will require sustained surveillance for a long period, given the low MTC incidence (one case per 2,299 person-years).

Importantly our data indicate that MTC onset in moderate-risk variant carriers occurs predominantly in adulthood, in genotype-first and even in phenotype-first approaches (with 100% and 84% developing MTC after age 30, respectively). This late-onset pattern may not meet the criteria for newborn genomic screening programmes (genotype first study design), which typically prioritise conditions where majority manifesting disease before age 5.^8^

Despite analysing nearly 500,000 individuals, we had limited numbers with *RET* pathogenic variants, particularly high-risk variants, highlighting the need for larger studies. We also lack all known pathogenic moderate risk variants in our study so it is possible that some may have different risk. Whilst appropriate for our research question, UK Biobank cohort show healthy recruitment effect as clearly observed in sex specific cancers.^31^ This and the minimum recruitment age of 40 years could have depleted the cohort of high/highest risk variants and missed MTC cases who died earlier. However, this potential depletion does not apply to incidentally identified *RET* variants in over 40 year olds and their follow up data. We observed a similar age distribution between variant carriers and non-carriers and similar prevalence of MTC in our cohort compared to previous nationwide estimates (7.5 vs 3.8 per 100,000) suggesting this might not have significantly impacted our findings.^26^ Our use of electronic health records to define MTC may have missed some cases, but our broad inclusion criteria encompassing any thyroid cancer, thyroidectomy (regardless of indication), and self-reported data likely captured most cases. We do not have calcitonin in our study cohort and incorporating this marker could provide more personalised risk estimates. We were unable to confirm the variant using a second DNA analysis method, although we mitigated against technical false positives through manual IGV checking. Moreover, all 52 variants that were subjected to Sanger sequencing at Geisinger (as part of MyCode Genome Screening and Counselling Program) had the presence of the variant was confirmed. The replication of findings across two different healthcare settings in two countries strengthens our conclusions, though due to all the limitations our estimates likely represent a lower bound of true population penetrance. While additional data on family members of clinically referred patients as a comparator group would have been useful, it’s absence does not affect our primary results.

In conclusion, we show that moderate-risk *RET* pathogenic variants predominate in incidental cases and carry substantially lower MTC risk than clinically ascertained cases, with no excess mortality without intervention. This evidence addresses a critical knowledge gap supporting informed clinical decision-making for individuals with incidentally identified *RET* variants.

## Acknowledgements

This research has been conducted using the UK Biobank Resource. This work was carried out under UK Biobank project number 103356. The current work is funded by the Wellcome Trust (219606/Z/19/Z) and summer studentship from Society of Endocrinology. The work is supported by the National Institute for Health Research (NIHR) Exeter Biomedical Research Centre, Exeter, UK. The Wellcome Trust and NIHR had no role in the design and conduct of the study; collection, management, analysis, and interpretation of the data; preparation, review, or approval of the manuscript; and decision to submit the manuscript for publication. The views expressed are those of the author(s) and not necessarily those of the Wellcome Trust, Department of Health, NHS or NIHR. For the purpose of open access, the author has applied a CC BY public copyright licence to any Author Accepted Manuscript version arising from this submission. The authors are grateful to Dr. Nicholas Purdy and Ms. Juliann Savatt for sharing the information of their study cohort^20^, which allows us to remove them from our survival analysis.

We are grateful to the MyCode participants for the use of their genomic and electronic health information, without whom this study would not possible. The patient enrolment and exome sequencing were funded by the Regeneron Genetics Centre. We thank the Geisinger-Regeneron DiscovEHR collaboration for making the genotype and phenotype data available.

## Data availability

All the UK Biobank data used in this study is freely accessible from the UK Biobank https://www.ukbiobank.ac.uk. The pathogenic variants used in the study are already available in manuscripts. Data from the Geisinger MyCode cohort is available through the MyCode Community Initiative and the Geisinger-Regeneron DiscovEHR collaboration.

