## Supplementary Data for "Mutation Spectrum and Associated Risks of Medullary Thyroid Cancer and All-Cause Mortality in Incidentally Identified MEN2A-Causing *RET* Variants"

**Supplementary Table 1: Characteristic of study Cohorts**

| | UK Biobank | Geisinger cohort | Exeter Clinical cohort |
|---|---|---|---|
| **N (unrelated)** | 383,914 | 122,640 | 1078 |
| **Cohort setting** | Population from the UK | Heath system based regional cohort, Pennsylvania, USA | Clinically referred cases from the UK for *RET* genetic testing for suspected MEN2 |
| **Age at recruitment, y** | 57.3 (8.11) | 59.1 (18.0) | 47.3 (18.0) |
| **Female Sex, n (%)** | 198,442 (51.7) | 73,602 (60.0) | 592 (56.4) |
| **Genetic data** | Whole exome | Whole exome | Targeted gene panel or Sanger sequencing |
| **Phenotype** | Electronic health record – Hospital, GP, cancer registry, surgical records, self-report | Electronic health record - Hospital, GP, cancer registry, surgical records | Clinician reported |

Data is given as n (%) for categorical variable and mean (SD) for the continuous variables

**Supplementary table 2:** Summary of the codes used to define both the strict and permissive definitions

| Definitions used in the study | Electronic health records codes (AND/OR) used in the definition (data combined from Self report, Hospital episode statistics or General practice records) |
|---|---|
| **1. Main definition: malignant thyroid cancer with medullary histology** | Thyroid Cancer: C73 (ICD10), 193/1939 (ICD9), 1065(Self-Reported), B53../BB9B./ByuB./ZV10y/X40Ia/44A2./X78cT/Xa98X/B9240/BB5fz/BB5f./BX78cV (GP records)<br>Histology in cancer registry: 8510 (Cancer Registry), X78cT (GP records) |
| **2. Broad definition: Definition 1 AND/OR Any thyroid cancer or thyroidectomy for any indication** | Thyroid Cancer: C73 (ICD10), 193/1939 (ICD9), 1065(Self-Reported), B53../BB9B./ByuB./ZV10y/X40Ia/44A2./X78cT/Xa98X/B9240/BB5fz/BB5f./BX78cV (GP records)<br>Thyroidectomy: B081/B082/B083/B084/B085/B086/B088/B089 (OPCS4), 71100/71102/71103/71104/71105/71101/71106/7110y/7110z/7110./711../71130/7113z/7113y/Xa80R/Xa80S/XE0AE/X40GP/XM0my (GP records) |

### Supplementary Table 3: MEN2-causing *RET* pathogenic variant classification and number in each study cohorts.

| HGVS Nomenclature CDNA | HGVS Nomenclature Protein | American Thyroid association classification | Present in UK Biobank unrelated*, | Present in Geisinger cohort unrelated, | Clinical Cohort unrelated, n | The American College of Medical Genetics and Genomics classification |
|---|---|---|---|---|---|---|
| **1597G>A** | Gly533Cys | Moderate | - | - | 1 | PS3 PP1 PM2 PP4 |
| **1826G>A or 1826G>T** | Cys609Tyr or Cys609Phe | Moderate | 7 | 2 | 5 | PP1 PS4 PM1 PM2 PP3 PP4 or PP1 PM2 PM1 PS4 PP4 PP3 |
| **1831T>C** | Cys611Arg | Moderate | - | - | 1 | PM5 PM2 PS3 |
| **1852T>G** | Cys618Gly | Moderate | - | - | 1 | PM1 PS4 PM2 PP3 PP4 |
| **1852T>C** | Cys618Arg | Moderate | - | - | 6 | PP1 PS4 PS3 PM1 PM2 PP4 |
| **1853G>C** | Cys618Ser | Moderate | - | - | 5 | PP1 PS1 PM5 PM2 PP3 PS4 |
| **1858T>G** | Cys620Gly | Moderate | - | 2 | 1 | PM5 PM2 PP4 PP3 |
| **1858T>C** | Cys620Arg | Moderate | - | - | 7 | PS4 PP1 PS3 PM1 PM2 PP4 |
| **1858T>A** | Cys620Ser | Moderate | - | - | 3 | PP1 PM1 PS4 PM2 PP3 PP4 |
| **1859G>T** | Cys620Phe | Moderate | - | - | 1 | PM5 PM2 PS4 PP4 PP3 |
| **1859G>A** | Cys620Tyr | Moderate | - | 3 | - | PM5 PM2 PS4 PP4 PP3 |
| **1860C>G** | Cys620Trp | Moderate | - | 1 | - | PM5 PM2 PS4 PP4 PP3 |
| **1996A>G** | Lys666Glu | Moderate | 6 | 2 | 2 | PS3 PM2 PP1 PP5 |
| **1998G>C** | Lys666Asn | Moderate | 7 | 1 | - | PS1 PM2 PM5 PS3 PS4 PP4 |
| **1998G>T** | Lys666Asn | Moderate | 7 | 5 | - | PS1 PM2 PM5 PS3 PS4 PP4 |
| **2018A>C** | Glu673Ala | Moderate | - | - | 1 | PS3 PP1 PP4 |
| **2304G>C or 2304G>T or2370G>C or 1891G>T or 2711C>T or 2752A>G** | Glu768Asn or Glu768Asp or Leu790Phe or Asp631Tyr or Ser904Phe or Met918Val | Moderate | 11 | 2 | - | PS3 PP1 PS4 PM2 PP4 or PS3 PP1 PS4 PM2 PP4 or PS1 PM2 PS4 PP1 PP3 PP4 or PS3 PP1 PS4 PM2 PP4 or PS3 PS4 PP1 PM2 PP4 or PM5 PP1 PM2 PP4 PP3 |
| **2370G>T** | Leu790Phe | Moderate | 11 | - | 7 | PS1 PM2 PS4 PP1 PP3 PP4 |
| **2410G>A** | Val804Met | Moderate | 95 | 30 | 14 | PS3 PP1 PP4 |
| **2410G>C** | Val804Leu | Moderate | 15 | - | - | PS1 PM5 PM2 PP4 PP3 PS3 PP1 PS4 |
| **2671T>G** | Ser891Ala | Moderate | 9 | 25 | 8 | PS3 PS4 PP1 PP4 |
| **1900T>C or 1900T>G or 1900T>A or 1901G>A or 1901G>T or 1902C>G or 2647_2648delinsTT** | Cys634Arg or Cys634Gly or Cys634Ser or Cys634Tyr or Cys634Phe or Cys634Trp or Ala883Phe | High | 1 | 4 | 35 | PS3 PS4 PP1 PM1 PM2 PP4 (same for all) |
| **2753T>C** | Met918Thr | Highest | - | - | 18 | PS3 PS2 PM6 PS4 PM2 PP4 |

*inline with UKB publication policy n<5 were combined

**Supplementary Table 4: Clinical features of *RET* pathogenic variant carriers with any thyroid cancer including MTC or thyroidectomy.**

| Individuals, n | Cohort | Variant | MTC, n | Thyroid cancer or Thyroidectomy | Indication for thyroidectomy |
|---|---|---|---|---|---|
| **6** | UK Biobank | Cys634Tyr/ Leu790Phe/ Val804Met | 3/6 | 6/6 | MTC -3 , Pluriglandular neoplasm-1, Thyrotoxicosis -1, Non-toxic Multinodular goitre -1 |
| **1** | Geisinger MyCode | Cys609Tyr | Yes | Yes | MTC |
| **1** | Geisinger MyCode | Cys620Gly | Yes | Yes | MTC |
| **1** | Geisinger MyCode | Cys620Gly | No | Yes | Prophylactic Thyroidectomy |
| **1** | Geisinger MyCode | Cys620Tyr | Yes | Yes | MTC |
| **1** | Geisinger MyCode | Cys620Tyr | Yes | Yes | MTC |
| **1** | Geisinger MyCode | Cys620Tyr | Yes | Yes | MTC |
| **1** | Geisinger MyCode | Cys634Arg | Yes | Yes | MTC |
| **1** | Geisinger MyCode | Cys634Phe | No | Yes | Unknown – Surgery not Performed at Geisinger |
| **1** | Geisinger MyCode | Val804Met | Yes | Yes | MTC |
| **1** | Geisinger MyCode | Val804Met | No | Yes | Hyperthyroidism |
| **1** | Geisinger MyCode | Ser891Ala | Yes | Yes | MTC |
| **1** | Geisinger MyCode | Ser891Ala | Yes | Yes | MTC |
| **1** | Geisinger MyCode | Ser891Ala | Yes | Yes | MTC |

*inline with UKB publication policy n<5 were combined

**Supplementary Table 5: Cases of Medullary Thyroid Cancer in study cohort and by American Thyroid Association pathogenic *RET* variant categories.** Prevalent and incident cases for UK Biobank and prevalent cases for Geisinger and clinical cohort.

| | **UK Biobank** | **Geisinger MyCode** | **Clinical cohort** |
|---|---|---|---|
| Whole cohort | 29/383914 (0.0075%) (7.5/100,000, CI 5.06 - 10.8) | 23/122640 (0.0187%) (18.7/100,000 CI 11.9-28.1) | 779/1078 (72.3%) |
| RET pathogenic variant carriers – ALL | 3/169 (1.8% CI 0.4-5.1) | 10/77 (12.9% CI 6.4-22.6) | 96/117 (82.1% CI 73.9-88.5) |
| Highest risk | - | - | 8/18 (44.4% CI 21.5-69.2) |
| High risk | 1/1 (100% CI 2.5-100) | 1/4 (25% CI 0.6-80.6) | 29/35 (82.9% CI 66.4-93.4) |
| Moderate risk* | 2/168 (1.2% CI 0.1-4.2) | 9/73 (12.3% CI 5.8-22.1) | 59/64 (92.2% CI 82.7-97.4) |
| p.(Val804Met)* | 1/95 (1.1% CI 0.02-5.7) | 1/30 (3.3% CI 0.1-17.2) | 13/14 (92.9% CI 66.1-99.8) |
| Moderate risk excluding p.(Val804Met)* | 1/73 (1.4% CI 0.03-7.4) | 8/43 (18.6% CI 8.4-33.4) | 46/50 (92.0% CI 82.8-99.9) |
| Moderate risk Extracellular variants* | 0/8 (0%) | 5/8 (62.5 CI 29.0-96.0) | 29/30 (96.7% CI 82.8-99.9) |

*MTC comparison in non p.(Val804Met) Vs p.(Val804Met) moderate risk carriers in UK biobank (fisher exact test p=1), MTC comparison in intracellular vs extracellular moderate risk carriers in UK biobank (fisher exact test p=1). MTC comparison in non p.(Val804Met) Vs p.(Val804Met) moderate risk carriers in Geisinger cohort (fisher exact test p=0.07), MTC comparison in intracellular vs extracellular moderate risk carriers in UK biobank (fisher exact test p=0.004).

**A)**

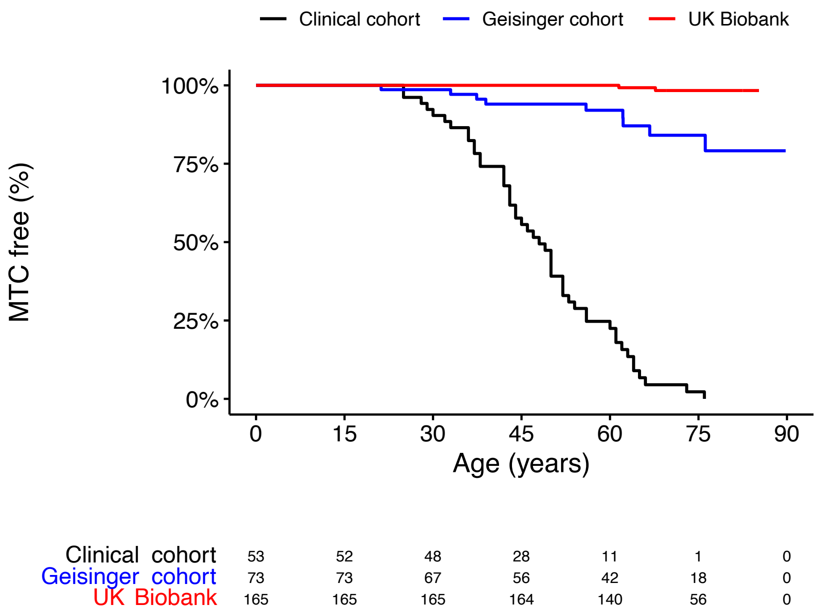

Clinical cohort
Geisinger cohort
UK Biobank
MTC free (%)
100%
75%
50%
25%
0%
0
15
30
45
60
75
90
Age (years)
Clinical cohort 53 52 48 28 11 1 0
Geisinger cohort 73 73 67 56 42 18 0
UK Biobank 165 165 165 164 140 56 0


*B)*

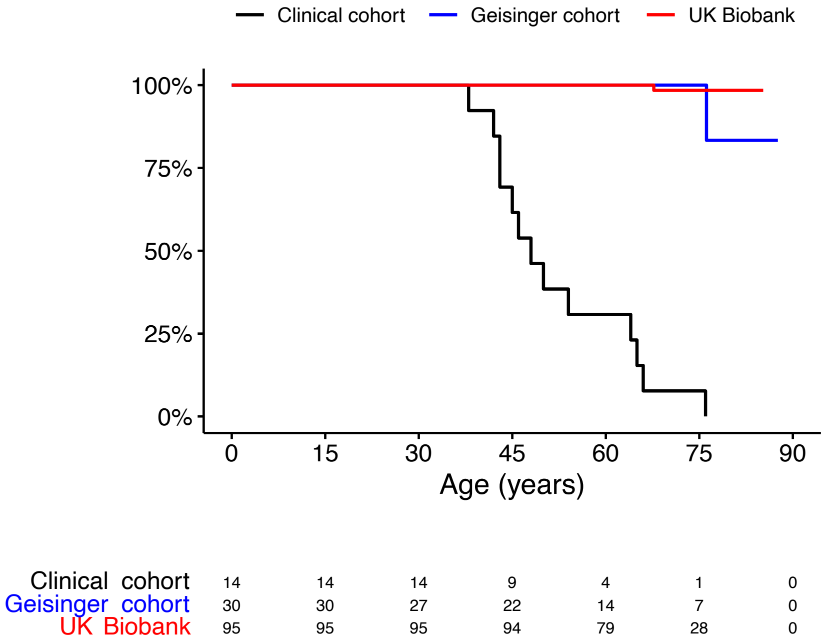

Clinical cohort
Geisinger cohort
UK Biobank
MTC free (%)
100%
75%
50%
25%
0%
0
15
30
45
60
75
90
Age (years)
Clinical cohort 14 14 14 9 4 1 0
Geisinger cohort 30 30 27 22 14 7 0
UK Biobank 95 95 95 94 79 28 0


*C)*

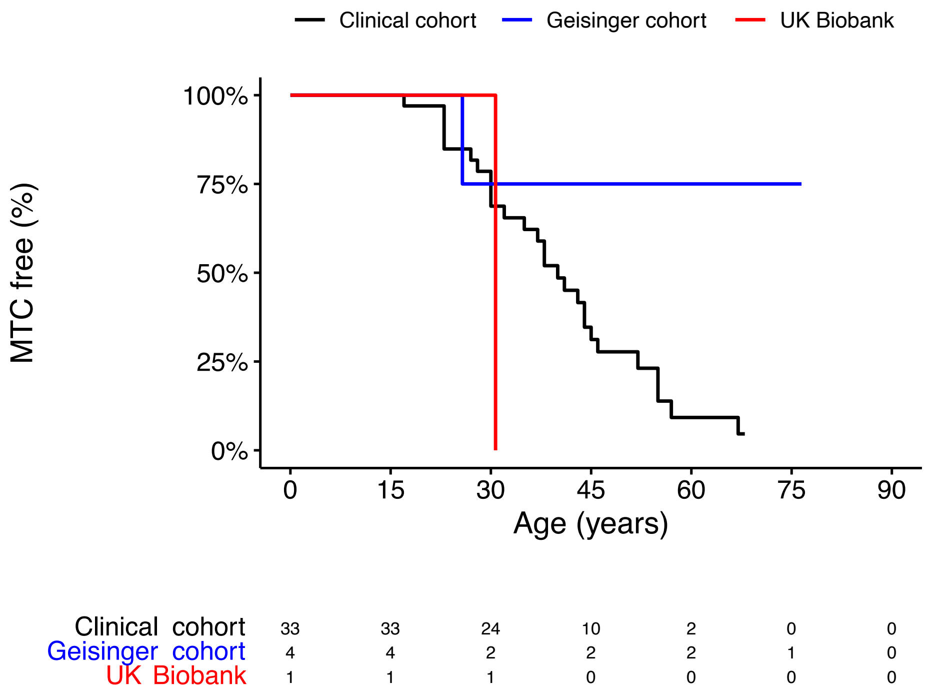


**Supplementary Figure 1: The age-related risk of medullary thyroid cancer in individuals with RET pathogenic variant ascertained clinically and in clinically unselected population and health system-based cohort with the matched variants.**

A) Kaplan-Meier plot demonstrating the age-related penetrance of MTC for RET moderate risk pathogenic variant carriers identified from individuals referred to the Exeter genomic laboratory for genetic testing in routine clinical practice in the UK with suspected MEN2A and unselected population cohort (UK Biobank) and health system-based cohort (Geisinger cohort). B) limiting to carriers with p.V804M in all three cohorts C) carriers with high-risk variants in study cohorts.
